# Metabolic Control and the Symptoms/Problems Domain of Kidney Disease-Specific Quality of Life in Patients Receiving Kidney Replacement Therapy

**DOI:** 10.64898/2026.10.16.26365777

**Authors:** David Moran

**Author notes:** Corresponding author: David Moran.

## Abstract

**Background:** Health-related quality of life (HRQoL) is an important outcome in patients with chronic kidney disease (CKD) receiving kidney replacement therapy (KRT), but the relationship between comprehensive metabolic control and the Symptoms/Problems domain of kidney disease-specific HRQoL remains uncertain.

**Objective:** To evaluate the association between composite metabolic control and the Symptoms/Problems domain of the Kidney Disease Quality of Life-36 (KDQOL-36) in patients with stage 5 CKD receiving KRT, and to explore the relationship between individual metabolic parameters and the continuous Symptoms/Problems score.

**Materials and Methods:** We conducted an observational, cross-sectional, analytical study of 177 adults receiving hemodialysis or peritoneal dialysis at a nephrology unit in Chihuahua, Mexico. Metabolic control included blood pressure, glycated hemoglobin (HbA1c), total cholesterol, and triglycerides. Adequate control was defined as achievement of at least three of four study-defined operational thresholds. The KDQOL-36 Symptoms/Problems score was analyzed as a continuous variable and categorized as low, moderate, or high according to the study protocol. Group comparisons, Spearman correlations, Pearson chi-square tests, and ordinal logistic regression were performed.

**Results:** Adequate metabolic control was present in 106 patients (59.9%). No, statistically significant association was observed between achievement of at least three of four study-defined operational thresholds and Symptoms/Problems score category (χ^2^=3.983; p=0.137; Cramer’s V=0.150). None of the individual metabolic parameters differed significantly across Symptoms/Problems score categories or showed significant correlations with the continuous score. In ordinal logistic regression, adequate metabolic control yielded an odds ratio of 1.81 (95% CI: 0.98-3.35; p=0.057) after adjustment for age, sex, time receiving KRT, and KRT modality.

**Conclusions:** No, statistically significant association was identified between the operational definition of metabolic control used in this study and the KDQOL-36 Symptoms/Problems domain in patients with stage 5 CKD receiving KRT. The findings support evaluating HRQoL multidimensionally and warrant confirmation in larger longitudinal studies.

**Categories:** Family/General Practice, Internal Medicine, Nephrology

## Introduction

Chronic kidney disease (CKD) is a major public health problem because of its substantial morbidity and mortality burden and its close association with cardiovascular disease and metabolic abnormalities [1]. Its prevalence has increased over recent decades and represents a growing burden on healthcare systems [2]. In advanced stages, disease progression may lead to the need for kidney replacement therapy (KRT), a situation associated with a high clinical and cardiovascular burden [1,3]. In Mexico, CKD is likewise an increasingly important health problem, and the growing demand for KRT represents a challenge for healthcare services[4]. CKD is strongly associated with cardiometabolic abnormalities such as diabetes mellitus, hypertension, dyslipidemia, and obesity. These conditions may coexist and jointly contribute to cardiovascular risk and kidney disease progression [5]. The cardiovascular-kidney-metabolic framework recognizes the interaction among these factors and underscores the importance of comprehensive management of patients with advanced kidney disease [3,6]. However, evidence regarding the combined impact of controlling these factors in patients receiving KRT remains limited.

In addition to traditional clinical outcomes, HRQoL is an important outcome in people with CKD [7]. Disease progression and the need for KRT can affect multiple dimensions of patients’ lives, including physical functioning, symptoms, illness perception, and overall well-being [8]. Quality of life is also a multifactorial phenomenon that may be influenced by sociodemographic characteristics, comorbidities, anemia, nutritional status, time on dialysis, and KRT modality. Studies conducted in different populations, including Mexican patients [9], have documented substantial impairment in quality of life among people receiving dialysis [10]. The KDQOL-36 has demonstrated acceptable psychometric properties for assessing kidney disease-specific HRQoL [11]. Generic and CKD-specific instruments have been developed to assess this outcome. The KDQOL-36 combines generic health measures with three kidney disease-specific scales: Symptoms/Problems of Kidney Disease, Effects of Kidney Disease, and Burden of Kidney Disease, together with physical and mental component summary measures derived from the SF-12 [11,12]. Its use in patients receiving dialysis enables assessment of the impact of CKD and its treatment from the patient’s perspective [13]. Although several studies have explored the relationship between individual metabolic parameters and quality of life [6], the results have been heterogeneous, and evidence regarding the effect of comprehensive metabolic control on kidney disease-specific symptoms and HRQoL in patients with CKD receiving KRT remains limited [14]. Determining whether the achievement of metabolic targets considered jointly is associated with the Symptoms/Problems domain could contribute to a better understanding of factors associated with this patient-reported outcome [9]. Therefore, the objective of the present study was to evaluate the association between metabolic control and the Symptoms/Problems domain of the KDQOL-36 in patients with CKD receiving KRT, as well as to explore the relationship between individual metabolic components and the continuous Symptoms/Problems score.

## Materials And Methods

This was an observational, cross-sectional analytical study incorporating a retrospective review of electronic clinical records. The study was conducted in adult patients with stage 5 chronic kidney disease (CKD) receiving kidney replacement therapy (KRT) at the Nephrology Department of Ambulatory Medical Care Unit (UMAA) No. 68 of the Instituto Mexicano del Seguro Social (IMSS), Chihuahua, Mexico. Data collection took place from September 30, 2024, to March 25, 2025. During this period, the KDQOL-36 questionnaire was administered directly to participants specifically for this study, while clinical and laboratory data were retrospectively extracted from electronic medical records using the most recent available routine laboratory results. A total of 177 participants constituted the final analytic sample.

### Sample size and sampling

The sample size was estimated to be a priority using the single-proportion formula *n* = *Z*^2^*pq*/*d*^2^, with a 95% confidence level (*Z* = 1.96), an expected proportion of 0.515 (*p* = 0.515), *q* = 1 − *p* = 0.485, and an absolute precision of 0.05. This yielded an estimated sample size of 384 participants. The proportion used for the calculation was prespecified in the approved study protocol. Non-probability convenience sampling was used. Patients were subsequently screened according to the prespecified eligibility criteria. After application of the inclusion and exclusion criteria and assessment of data completeness, 177 participants constituted the final analytic sample.

### Clinical and laboratory measurements

Clinical and laboratory information was obtained retrospectively from the patients’ electronic clinical records. For each participant, the most recent available laboratory results documented in the medical record were used. The laboratory variables evaluated in the present study were glycated hemoglobin (HbA1c), total cholesterol, and triglycerides. Blood pressure measurements and relevant clinical characteristics were also obtained from the clinical record.

No blood or urine specimens were collected specifically for this research study. The investigators did not perform additional specimen processing or laboratory analyses. All laboratory values analyzed in this study corresponded to routine clinical testing performed as part of the patients’ usual medical care, and the study used the results documented in the institutional electronic clinical records. Urinary laboratory measurements were not included among the study variables, and no urine samples were collected or analyzed for research purposes. Blood pressure was considered controlled when systolic and diastolic blood pressure were ≤140/90 mmHg and uncontrolled when values were >140/90 mmHg. HbA1c was considered controlled when ≤6.5% and uncontrolled when >6.5%. Total cholesterol was considered controlled when 150 mg/dL. Because optimal HbA1c targets in dialysis-dependent kidney failure are not firmly established and HbA1c may have reduced reliability in this population, the HbA1c threshold was treated as a prespecified operational criterion rather than a universal treatment target [15].

### Composite metabolic control

A composite metabolic control variable was constructed from the four prespecified components: blood pressure, HbA1c, total cholesterol, and triglycerides. Metabolic control was considered adequate when the participant met at least three of the four prespecified operational thresholds and inadequate when two or fewer thresholds were met. As a sensitivity analysis, a more stringent definition requiring achievement of all four thresholds was also evaluated.

### Health-related quality-of-life assessment

Health-related quality of life was assessed using the Kidney Disease Quality of Life-36 (KDQOL-36). The standardized scoring procedure of the instrument was used to obtain domain scores [16]. For the present study, the 12-item Symptoms/Problems of Kidney Disease domain was selected as the primary outcome. The score is transformed to a 0-100 scale, with higher scores indicating fewer kidney disease-related symptoms/problems and better kidney disease-specific health-related quality of life. The continuous score was used for correlation analyses and was categorized according to the prespecified study protocol as low (80 points) for comparative analyses and ordinal logistic regression.

### Statistical analysis

The distribution of quantitative variables was assessed using the Shapiro-Wilk test. Quantitative variables were summarized as mean ± standard deviation or median and interquartile range, as appropriate, and categorical variables were summarized as absolute frequencies and percentages. HbA1c, total cholesterol, triglyceride concentrations, and blood pressure measurements were compared across the three Symptoms/Problems score categories using the Kruskal-Wallis test. Spearman correlation coefficients (ρ) were calculated between the continuous Symptoms/Problems score and the metabolic parameters. The association between categorized metabolic control and Symptoms/Problems score category was assessed using Pearson’s chi-square test, and the magnitude of the association was estimated using Cramer’s V. Ordinal logistic regression was performed using the Symptoms/Problems score category as the ordinal dependent variable. Univariable ordinal logistic regression models were fitted separately for metabolic control, age, sex, time receiving KRT, and KRT modality. A multivariable model included metabolic control, age, sex, time receiving KRT, and KRT modality. Results were expressed as odds ratios (ORs), 95% confidence intervals (95% CIs), and p-values.

### Ethics approval and informed consent

The research protocol was reviewed and approved by the Local Health Research Committee No. 805, Family Medicine Unit No. 33, Instituto Mexicano del Seguro Social (IMSS), Chihuahua, Mexico (Institutional Registration No. R-2024-805-026; approval date: September 23, 2024). Written informed consent was obtained from all participants. Confidentiality was maintained, and identifying information was excluded from the study database.

#### Payment/services info

The author have declared that no financial support was received from any organization for the submitted work.

#### Financial relationships

The author have declared that they have no financial relationships at present or within the previous three years with any organizations that might have an interest in the submitted work

## Data Availability Statement

The dataset generated and analyzed during the current study is available from the corresponding author upon reasonable request, subject to institutional authorization and applicable patient privacy and data-protection requirements. During manuscript preparation, ChatGPT (OpenAI) was used to translate the manuscript from Spanish into English and to assist with language editing, formatting, and editorial organization. All AI-assisted content was reviewed by the author, who remains fully responsible for the accuracy, integrity, and final content of the manuscript.

## Results

A total of 177 patients with CKD receiving KRT and treated at UMAA No. 68 of the IMSS were included. Mean age was 55 ± 10 years; 108 patients (61.0%) were men and 69 (39.0%) were women. Hemodialysis was performed in 83 patients (46.9%) and peritoneal dialysis in 94 (53.1%). Type 2 diabetes mellitus was present in 55.9% of patients, while systemic hypertension was the most common comorbidity (86.4%). Dyslipidemia was present in 28.8% of patients. The Symptoms/Problems score categories showed a similar distribution across KRT modalities. Among patients receiving peritoneal dialysis, 25.5% were low, 57.4% moderate, and 17.0% high Symptoms/Problems scores, whereas among those receiving hemodialysis the corresponding proportions were 22.9%, 65.1%, and 12.0%, respectively. No, statistically significant association was identified between KRT modality and Symptoms/Problems score category (χ^2^(2)=1.287; p=0.525; Cramer’s V=0.085) (Table 1).

**TABLE 1:** KRT modality and KDQOL-36 Symptoms/Problems score category’. Pearson’s chi-square test: χ^2^(2)=1.287; p=0.52S; Cramer’s V=0.085. Data are presented as n (%). Percentages arc row percentages.

| KRT modality | Low Symptoms/Problems score n (%) | Moderate Symptoms/Problems score n (%) | High Symptoms/Problems score n (%) | Total |
| --- | --- | --- | --- | --- |
| Peritoneal dialysis | 24 (25.5%) | 54 (57.4%) | 16 (17.0%) | 94 |
| Hemodialysis | 19 (22.9%) | 54 (65.1%) | 10 (12.0%) | 83 |
| Total | 43 (24.3%) | 108 (61.0%) | 26 (14.7%) | 177 |

Regarding metabolic control, 106 patients (59.9%) met at least three of the four study-defined operational thresholds and were classified as having adequate metabolic control, whereas 71 (40.1%) had inadequate metabolic control. Among the individual components, HbA1c had the highest proportion of patients meeting the study-defined threshold (79.1%), followed by total cholesterol (71.2%) and blood pressure (60.5%). Triglycerides had the lowest proportion of patients meeting the study-defined threshold, with 50.8% classified as controlled (Table 2).

**TABLE 2:** Metabolic control and achievement of individual metabolic thresholds (n=177) Data are presented as n (%).

| Variable | Controlled n (%) | Uncontrolled n (%) |
| --- | --- | --- |
| **HbA1c** | **140 (79.1%)** | **37 (20.9%)** |
| Total cholesterol | 126 (71.2%) | 51 (28.8%) |
| Triglycerides | 90 (50.8%) | 87 (49.2%) |
| Blood pressure | 107 (60.5%) | 70 (39.5%) |
| Metabolic control $\geq 3$ of 4 thresholds | 106 (59.9%) | 71 (40.1%) |

No, statistically significant differences were observed in metabolic parameters across the Symptoms/Problems score categories. The median HbA1c was 6.0% in the low, moderate, and high Symptoms/Problems score groups (H=0.411; p=0.814). Similarly, no significant differences were observed in systolic blood pressure (H=0.835; p=0.659), diastolic blood pressure (H=0.037; p=0.982), total cholesterol (H=3.201; p=0.202), or triglycerides (H=1.913; p=0.384) (Table 3).

**TABLE 3:**
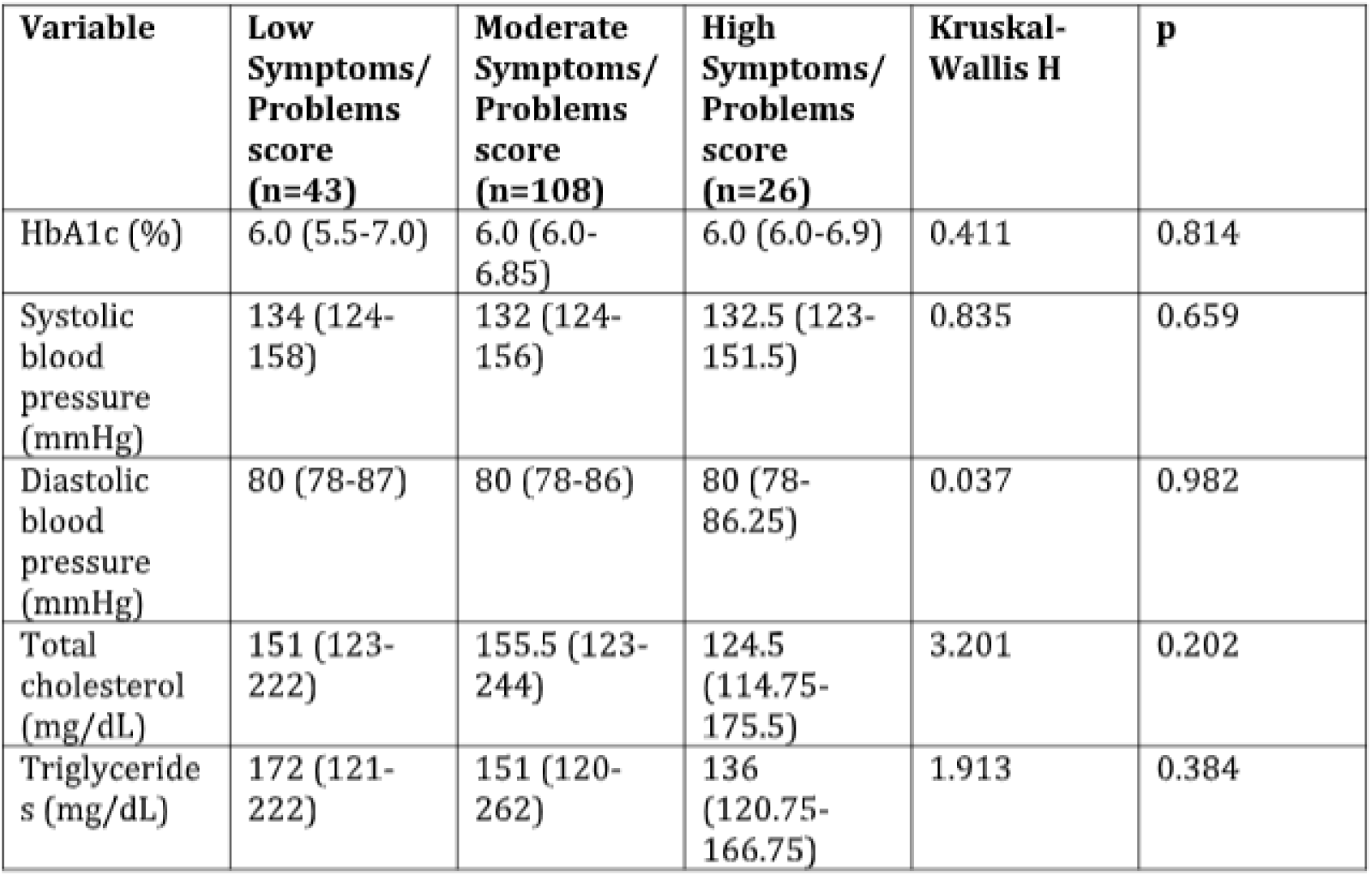
Metabolic parameters according to KDQOL-36 Symptoms/Problems score category (n=177) Data are presented as median (lower quartile-upper quartile). H denotes the Kruskal-Wallis statistic.

When the association between metabolic control and Symptoms/Problems score category was analyzed, 23 patients (21.7%) with adequate metabolic control had low, 63 (59.4%) had moderate, and 20 (18.9%) had high Symptoms/Problems scores. Among patients with inadequate metabolic control, 20 (28.2%) had low, 45 (63.4%) had moderate, and 6 (8.5%) had high Symptoms/Problems scores. The association between achievement of at least three of the four study-defined operational thresholds and Symptoms/Problems score category was not statistically significant (χ^2^=3.983; p=0.137; Cramér’s V=0.150) (Table 4).

**TABLE 4:**
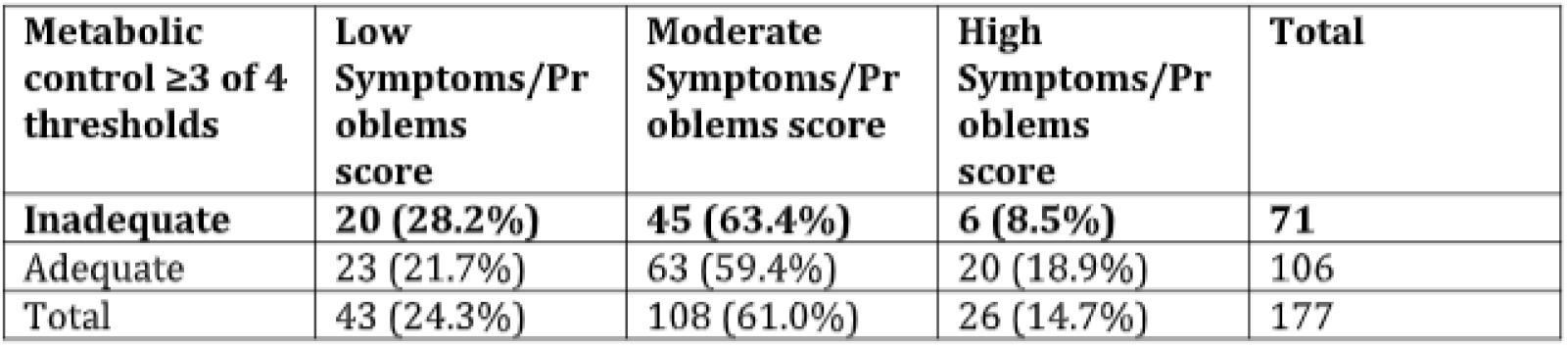
Association between metabolic control (≥3 of 4 thresholds) and KDQOI.-36 Symptoms/Problems score category. Data are presented as n (%). Percentages are row percentages. Pearson’s chi-square test: X^2^(2)=3.983; p=0.137; Cramer’s V=0.150.

No, statistically significant correlations were identified between the continuous Symptoms/Problems score and any of the metabolic parameters evaluated (Table 5). Correlations were weak and close to zero for HbA1c (ρ=−0.001; p=0.988), systolic blood pressure (ρ=−0.105; p=0.165), diastolic blood pressure (ρ=0.011; p=0.888), total cholesterol (ρ=−0.082; p=0.278), and triglycerides (ρ=−0.034; p=0.651).

**TABLE 5:** Spearman correlations between KDQOL-36 Symptoms/problems score and metabolic parameters (n=177) Data are Spearman correlation coefficients (ρ) between the continuous KDQOL-36 Symptoms/Problems score and each metabolic parameter.

| Metabolic parameter | Spearman's $\rho$ | p |
| --- | --- | --- |
| HbA1c (%) | -0.001 | 0.988 |
| Systolic blood pressure (mmHg) | -0.105 | 0.165 |
| Diastolic blood pressure (mmHg) | 0.011 | 0.888 |
| Total cholesterol (mg/dL) | -0.082 | 0.278 |
| Triglycerides (mg/dL) | -0.034 | 0.651 |

In univariable ordinal logistic regression analyses, adequate metabolic control was not significantly associated with Symptoms/Problems score category (OR=1.70; 95% CI: 0.93-3.13; p=0.085). After adjustment for age, sex, time receiving KRT, and KRT modality, the association remained non-significant (OR=1.81; 95% CI: 0.98-3.35; p=0.057) (Table 6).

**TABLE 6:** Ordinal logistic regression: composite metabolic control and KDQOL-36 Symptoms/Problems score category (n=177) OR. odds ratio; Cl, confidence interval: KRT. kidney replacement therapy. Univariable ordinal logistic regression analyses were performed separately for each predictor. The adjusted model Included metabolic control, age, sex, time receiving KRT, and KRT modality. Inadequate metabolic control, female sex, and peritoneal dialysis were the reference categories. Age and time receiving KRT were modeled per year. All regression analyses included 177 participants.

| Predictor | Univariable model OR (95% CI) | p | Adjusted model OR (95% CI) | p |
| --- | --- | --- | --- | --- |
| Adequate metabolic control vs inadequate | 1.70 (0.93-3.13) | 0.085 | 1.81 (0.98-3.35) | 0.057 |
| Age, per year | 0.98 (0.96-1.00) | 0.093 | 0.98 (0.95-1.00) | 0.065 |
| Time receiving KRT, per year | 0.99 (0.89-1.10) | 0.852 | 0.99 (0.89-1.10) | 0.864 |
| Male vs female | 1.24 (0.68-2.26) | 0.490 | 1.10 (0.60-2.03) | 0.752 |
| Hemodialysis vs peritoneal dialysis | 0.94 (0.52-1.70) | 0.842 | 0.91 (0.50-1.64) | 0.752 |

In a complementary multivariable ordinal logistic regression model incorporating the metabolic parameters as continuous variables, no statistically significant associations with Symptoms/Problems score category were identified (Table 7).

**TABLE 7:**
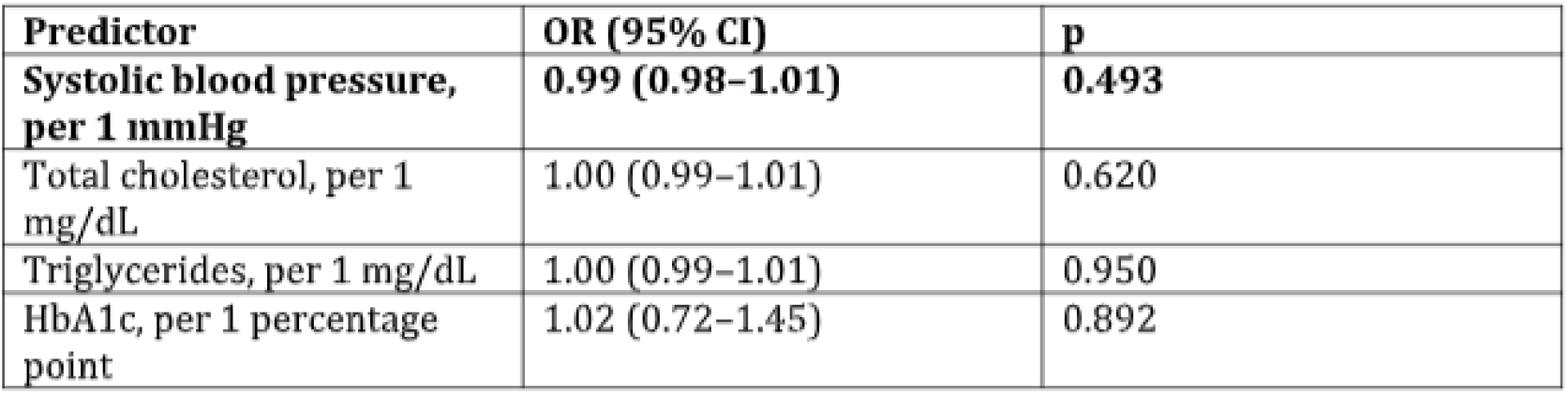
Multivariable ordinal logistic regression with continuous metabolic markers and KDQOL-36 Symptoms/Problems score category (n= 177) OR, odds ratio; Cl, confidence interval. A multivariable ordinal logistic regression model was fitted with all four continuous metabolic markers entered simultaneously. Systolic blood pressure was modeled per 1 mmHg, total cholesterol per 1 mg/dL, triglycerides per 1 mg/dL, and HbA1c per 1 percentage point The analysis included 177 participants. Overall model: χ^2^(4)=1.720; p=0.787; Nagelkerke R^2^=0.011; test of parallel lines p=0.657.

| Predictor | OR (95% CI) | p |
| --- | --- | --- |
| Systolic blood pressure, per 1 mmHg | 0.99 (0.98–1.01) | 0.493 |
| Total cholesterol, per 1 mg/dL | 1.00 (0.99–1.01) | 0.620 |
| Triglycerides, per 1 mg/dL | 1.00 (0.99–1.01) | 0.950 |
| HbA1c, per 1 percentage point | 1.02 (0.72–1.45) | 0.892 |

As a sensitivity analysis, a more stringent definition of metabolic control was used, requiring achievement of all four thresholds. The results were consistent with the primary analysis, and no statistically significant association was identified between complete metabolic control and Symptoms/Problems score category (χ^2^(2)=3.413; p=0.182; Cramer’s V=0.139).

## Discussion

In this study of patients with CKD receiving KRT, no statistically significant association was identified between the operational definition of metabolic control and the Symptoms/Problems domain of kidney disease-specific HRQoL. This finding was consistent across comparison of Symptoms/Problems score categories, correlation analyses using the continuous score, and ordinal logistic regression. The adjusted model produced an OR above one (1.81), but the 95% CI included the null value (0.98-3.35), and the p-value was 0.057, so the association was not statistically significant.

Our findings are consistent with part of the available evidence, although the literature shows heterogeneous results regarding metabolic factors and quality of life. Afsar et al. found no significant association between metabolic syndrome and quality of life or mood in patients receiving hemodialysis [17]. In contrast, Masoumi et al. reported associations of metabolic syndrome with age, physical activity, mood disturbances, and sleep quality in a hemodialysis population [18]. These findings support a multifactorial view of quality of life in KRT, in which metabolic factors interact with nutritional, inflammatory, clinical, behavioral, and social factors. Babovic et al. also identified biochemical, immunonutritional, and social-support factors associated with quality of life in patients receiving hemodialysis [14].

A multifactorial explanation is also supported by longitudinal and observational studies. Quality of life may change during kidney disease and treatment [19,20], including during transition to dialysis or transplantation [21]. Shumbusho et al. identified associations between KDQOL-36 scores and sex, employment, time on hemodialysis, hemoglobin concentration, and comorbidity burden [8]. Dehesa-López et al. found clinical and dialysis-related factors associated with poorer HRQoL in Mexican patients on hemodialysis [9]. Van Haalen et al. reported associations between hemoglobin levels and different dimensions of HRQoL in CKD, while Hoshino et al. described relationships between hemoglobin, HRQoL, and physical activity in stage 3-5 non-dialysis CKD [22,23]. The latter population differs from ours because it did not include patients receiving dialysis, which limits direct comparability. These studies also evaluated broader or different HRQoL dimensions, whereas the present study focused specifically on the KDQOL-36 Symptoms/Problems domain.

The point estimate for adequate metabolic control was above one in both the unadjusted and adjusted ordinal models, but the confidence intervals crossed the null value. This consistency in direction should be interpreted cautiously because the study was not powered to establish a small association and the crosssectional design precludes assessment of temporality. Larger studies with longitudinal follow-up could determine whether composite metabolic control is related to subsequent changes in Symptoms/Problems scores.

KRT modality was not significantly associated with Symptoms/Problems score category in our population. This differs from some previous reports. Sanchez et al., in a cross-sectional study of 530 patients receiving different KRT modalities, reported differences in KDQOL-36 scores according to modality and association with comorbidity and socioeconomic factors [24]. Systematic review and meta-analysis also found differences between hemodialysis and peritoneal dialysis in some HRQoL domains, without demonstrating uniform superiority of one modality [25]. Other studies likewise report heterogeneous findings across modalities [26,27]. Differences in patient characteristics, treatment context, and socioeconomic conditions may contribute to this variability.

The use of HbA1c as one component of the composite metabolic definition also warrants caution in patients receiving dialysis. Current evidence indicates that kidney failure-related factors can reduce the reliability of HbA1c and that optimal glycemic targets for the dialysis population remain uncertain; therefore, our HbA1c threshold should be interpreted as a study-specific operational criterion rather than a universal treatment target [15].

None of the individual metabolic parameters showed a statistically significant association with the Symptoms/Problems domain in our population. No differences were observed in HbA1c, blood pressure, total cholesterol, or triglycerides across Symptoms/Problems score categories, and correlations with the continuous score were weak and close to zero. The continuous-variable ordinal model yielded the same overall interpretation. In patients with diabetes and kidney failure, interventions targeting self-care and glycemic management have produced improvements in HbA1c and selected quality-of-life dimensions, but these findings require confirmation in larger studies [28].

The sensitivity analysis using the stricter four-of-four definition produced a similar result (χ^2^(2)=3.413; p=0.182; Cramer’s V=0.139), suggesting that the primary finding was not dependent on the specific composite-control definition. The study has several strengths, including the inclusion of 177 patients receiving different KRT modalities, use of a kidney disease-specific quality-of-life instrument and its standardized scoring procedure, assessment of metabolic control as both a composite and individual components, and evaluation through complementary statistical approaches. Limitations include the crosssectional, single-center design; the inability to establish temporality or causality; limited generalizability; and the absence of potentially important nutritional, hematologic, psychological, and social covariates. In addition, the present analysis focused on the Symptoms/Problems domain rather than the full multidomain KDQOL-36 instrument; therefore, the findings should not be interpreted as evidence regarding all dimensions of overall HRQoL. The sample size may also have limited detection of small associations.

## Conclusions

Among patients with stage 5 CKD receiving KRT, no statistically significant association was identified between the operational definition of metabolic control used in this study and the Symptoms/Problems domain of the KDQOL-36. This finding remained consistent when metabolic control was evaluated as a composite variable, through its individual components, and across complementary statistical analyses. These results support a multidimensional approach to HRQoL assessment, recognizing that the Symptoms/Problems domain represents only one component of the broader patient-reported impact of kidney disease.

## References

1. Kalantar-Zadeh K, Jafar TH, Nitsch D, Neuen BL, Perkovic V: Chronic kidney disease . Lancet. 2021, 398:786–802. 10.1016/S0140-6736(21)00519-5

2. Junejo S, Chen M, Ali MU, Ratnam S, Malhotra D, Gong R: Evolution of chronic kidney disease in different regions of the world. J Clin Med. 2025, 14:4144. 10.3390/jcm14124144

3. Rangaswami J, Shlipak MG, Mathew RO, Ndumele CE: The cardiovascular-kidney-metabolic health framework: implications for nephrology. Clin J Am Soc Nephrol. 2025, 20:1299–1306. 10.2215/CJN.0000000744

4. Argaiz ER, Morales-Juárez L, Razo C, Ong L, Rafferty Q, Rincón-Pedrero R, Gamba G: La carga de enfermedad renal crónica en México. Análisis de datos basado en el estudio Global Burden of. Disease. 20212023, 159:501–508. 10.24875/GMM.23000393

5. Zhu D, Judge PK, Wanner C, Haynes R, Herrington WG: The prevention and management of chronic kidney disease among patients with metabolic syndrome. Kidney Int. 2025, 107:816–824. 10.1016/j.kint.2024.12.021

6. Wright WL, Urquhart S, Brunton S: Beyond blood glucose and blood pressure control in type 2 diabetes: alternative management strategies to prevent the development and progression of CKD. J Prim Care Community Health. 2023, 14:21501319231153599. 10.1177/21501319231153599

7. Ware JE Jr, Richardson MM, Meyer KB, Gandek B: Improving CKD-specific patient-reported measures of health-related quality of life. J Am Soc Nephrol. 2019, 30:664–677. 10.1681/ASN.2018080814

8. Shumbusho G, Hategeka C, Vidler M, Kabahizi J, McKnight M: Health-related quality of life of patients undergoing in-centre hemodialysis in Rwanda: a cross-sectional study. BMC Nephrol. 2022, 23:345. 10.1186/s12882-022-02958-6

9. Dehesa-López E, Correa-Rotter R, Olvera-Castillo D, González-Parra C, Baizabal-Olarte R: Clinical, dialytic, and laboratory factors associated with poor health-related quality of life in Mexican patients on hemodialysis. Rev Invest Clin. 2016, 68:192–200.

10. Luna-Ríos OA, Juárez-Sánchez JO: Calidad de vida en pacientes en terapia de sustitución renal en hemodiálisis. Rev Med Inst Mex Seguro Soc. 202361, 295:300.

11. Peipert JD, Bentler PM, Klicko K, Hays RD: Psychometric properties of the Kidney Disease Quality of Life 36-item short-form survey (KDQOL-36) in the United States. Am J Kidney Dis. 2018, 71:461–468. 10.1053/j.ajkd.2017.07.020

12. Yang F, Wong CKH, Luo N, Piercy J, Moon R, Jackson J: Mapping the Kidney Disease Quality of Life 36-item short form survey (KDQOL-36) to the EQ-5D-3L and the EQ-5D-5L in patients undergoing dialysis. Eur J Health Econ. 2019, 20:1195–1206. 10.1007/s10198-019-01088-5

13. Hays RD, Kallich JD, Mapes DL, Coons SJ, Carter WB: Development of the Kidney Disease Quality of Life (KDQOL) instrument. Qual Life Res. 1994, 3:329–338. 10.1007/BF00451725

14. Babovic B, Belada Babovic N, Tomovic F, Radovanovic S, Debeljevic M, Mustur D, Mihaljevic O: The importance of biochemical parameters, immunonutritional status, and social support for quality of life in chronic hemodialysis patients. Medicina (Kaunas. 2024, 60:1751. 10.3390/medicina60111751

15. Klein KR, Lingvay I, Tuttle KR, Flythe JE: Glycemic management and individualized diabetes care in dialysis-dependent kidney failure. Diabetes Care. 2025, 48:164–176. 10.2337/dci24-0081

16. Kidney Disease Quality of Life Working Group: The KDQOL-36™ Survey. RAND Health Care. Available from: https://www.rand.org/health/surveys_tools/kdqol.html.

17. Afsar B, Elsurer R, Sezer S, Ozdemir NF: Does metabolic syndrome have an impact on the quality of life and mood of hemodialysis patients?. J Ren Nutr. 2009, 19:365–371. 10.1053/j.jrn.2009.01.016

18. Masoumi M, Shahidi M, Montazeri SMH, et al.: The prevalence and risk factors of metabolic syndrome in patients with hemodialysis. J Caring Sci. 2024, 13:27–35. 10.34172/jcs.2024.32974

19. Mujais SK, Story K, Brouillette J, et al.: Health-related quality of life in CKD patients: correlates and evolution over time. Clin J Am Soc Nephrol. 2009, 4:1293–1301. 10.2215/CJN.05541008

20. Wu AW, Fink NE, Marsh-Manzi JV, Meyer KB, Finkelstein FO, Chapman MM, Powe NR: Changes in quality of life during hemodialysis and peritoneal dialysis treatment: generic and disease-specific measures. J Am Soc Nephrol. 2004, 15:743–753. 10.1097/01.asn.0000113315.81448.ca

21. Hakamäki M, Järvisalo MJ, Lankinen R, et al.: Evolution of quality of life in chronic kidney disease stage 4-5 patients transitioning to dialysis and transplantation. Nephron. 2022, 146:439–448. 10.1159/000521771

22. Van Haalen H, Jackson J, Spinowitz B, Milligan G, Moon R: Impact of chronic kidney disease and anemia on health-related quality of life and work productivity: analysis of multinational real-world data. BMC Nephrol. 2020, 21:88. 10.1186/s12882-020-01746-4

23. Hoshino J, Muenz D, Zee J, et al.: Associations of hemoglobin levels with health-related quality of life, physical activity, and clinical outcomes in persons with stage 3-5 nondialysis CKD. J Ren Nutr. 2020, 30:404–414. 10.1053/j.jrn.2019.11.003

24. Sanchez S, Teelucksingh S, Ali R, Bailey H, Legall G: Quality of life and health status among patients receiving renal replacement therapy in Trinidad and Tobago, West Indies. Int J Nephrol Renovasc Dis. 2021, 14:173–192. 10.2147/IJNRD.S302157

25. Chuasuwan A, Pooripussarakul S, Thakkinstian A, Ingsathit A, Pattanaprateep O: Comparisons of quality of life between patients underwent peritoneal dialysis and hemodialysis: a systematic review and metaanalysis. Health Qual Life Outcomes. 2020, 18:191. 10.1186/s12955-020-01449-2

26. Alshehri M, Alshehri A, Alfageeh A, et al.: Who have a better-kidney-related quality of life: peritoneal dialysis or hemodialysis patients? A cross-sectional study from Saudi Arabia. BMC Nephrol. 2023, 24:216. 10.1186/s12882-023-03270-7

27. Yonata A, Islamy N, Taruna A, Pura L: Factors affecting quality of life in hemodialysis patients . Int J Gen Med. 2022, 15:7173–7178. 10.2147/IJGM.S375994

28. Griva K, Rajeswari M, Nandakumar M, et al.: The combined diabetes and renal control trial (C-DIRECT): a feasibility randomised controlled trial to evaluate outcomes in multi-morbid patients with diabetes and on dialysis using a mixed methods approach. BMC Nephrol. 2019, 20:2. 10.1186/s12882-018-1183-z

